# CLINICAL CHARACTERISTICS AND OUTCOMES OF HOSPITALIZED PATIENTS WITH HEART FAILURE AND SARCOIDOSIS: ANALYSIS OF THE NATIONWIDE READMISSIONS DATABASE 2010-2019

**DOI:** 10.1101/2023.08.25.23294650

**Authors:** Raheel Ahmed, Hiroyuki Sawatari, Khadija Amanullah, Joseph Okafor, Syed Emir Irfan Wafa, Saurabh Deshpande, Kamleshun Ramphul, Isma Ali, Mohammed Khanji, Athol Wells, Omar Ezzeddin, Vasilis Kouranos, Rakesh Sharma, Virend K. Somers, Selma F. Mohammed, C. Anwar A. Chahal

## Abstract

**BACKGROUND:** Sarcoidosis is a multi-systemic granulomatous inflammatory disorder. In the setting of cardiac involvement, clinical manifestations include ventricular arrhythmias, high-grade atrioventricular block (AVB) and heart failure (HF). The impact of HF in patients with sarcoidosis has not been established from real-world data.

**METHODS:** Patients admitted with sarcoidosis from 2010-2019 were identified from the Nationwide Readmissions Database. Those with ischemic heart disease were excluded. Sarcoidosis patients without HF were propensity matched for age, gender and Charlson comorbidity index and compared to patients with HF. Clinical characteristics, length of stay (LOS), adjusted healthcare-associated costs (HAC), 90-day readmission and 90-day mortality was observed.

**RESULTS:** During the 10-year study period, 97,961 patients (median age 63 [54-71] years, 37.9% male) with a diagnosis of sarcoidosis were hospitalized (35.9% with HF and 64.1% without HF). On index admission, HF patients had a higher prevalence of AVB (3.3% vs 1.4%, p<0.0001), ventricular tachycardia (6.5% vs 1.3%, p<0.0001), ventricular fibrillation (0.4% vs 0.1%, p<0.0001) and atrial fibrillation (22.1% vs 7.5%, p<0.0001). The median LOS (4 [3-7] vs. 4[2-6] days, p<0.0001) was similar but median HAC (US$ 9,954.5 [5,934.7-18,128.8] vs. 8,828.3 [5,303.1-15,384.9], p<0.0001) during the index admission were higher in HF patients.

The LOS and HAC were greater in HF patients on 90-day readmission. HF patients were significantly more likely to be re-admitted within 90 days [adjusted all-cause readmissions (HR [95% CI: 1.28 [1.25 – 1.31], p<0.0001), atrial fibrillation (HR 1.35 [1.05-1.75], p=0.02), acute HF (HR 10.77 [9.45 – 12.16], p<0.0001) and ventricular tachycardia/ventricular fibrillation (HR 2.55 [1.69 – 3.85], p<0.0001)]. Adjusted inpatient mortality at readmission was also higher in HF patients (5.1% vs. 3.8%, p<0.0001).

**CONCLUSION:** The presence of HF in hospitalized sarcoidosis patients is associated with an increased prevalence of conduction disorders, ventricular arrhythmias and atrial fibrillation. HF patients had greater costs, readmissions and mortality at 90-days.

**What is known?:**

1) Cardiac involvement in sarcoidosis is associated with ventricular arrhythmias, high-grade atrioventricular block and heart failure
2) Retrospective small, single-center studies have reported relatively poor long-term survival outcomes for symptomatic cardiac sarcoidosis patients with reduced left ventricular ejection fraction

**What the study adds?:**

1) Using a large real-world database, this study has demonstrated that heart failure in hospitalized sarcoidosis patients is associated with increased prevalence of arrythmia, conduction disorders, cardiac implanted electronic devices, catheter ablations and cardiac transplantation.
2) Heart failure in hospitalized sarcoidosis patients leads to a significantly higher length of stay, healthcare-adjusted costs, 90-days readmissions and mortality following readmission.

## INTRODUCTION

Sarcoidosis is a multisystem inflammatory disorder characterized by the presence of non-necrotizing granulomas(1). The annual incidence ranges from 2.3 to 11 cases per 100,000 individuals with the majority occurring between the ages of 25 and 40(2, 3). Cardiac involvement is present in 5-10% of cases, although autopsy data suggests a higher prevalence approximating 20-30%(4). In the absence of an endomyocardial biopsy demonstrating non-caseating granuloma, a clinical diagnosis of cardiac sarcoidosis (CS) can be made using diagnostic criteria established by guidelines such as the Japanese Ministry of Health and Welfare and the 2014 Heart Rhythm Society Consensus (HRS) document(5, 6). The prevalence of CS varies according to region and is reported as 1:100,000 in South Korea and Taiwan, 3.7:100,000 in Japan, and 150:100000 in Sweden and Canada(7–12). When present, clinical manifestations include high-grade atrioventricular block (AVB), ventricular arrhythmias, heart failure (HF) and sudden cardiac death (SCD)(13–16).

Historically, studies have reported relatively poor long-term survival outcomes for symptomatic CS patients with reduced left ventricular ejection fraction (LVEF), an independent predictor of adverse events(17, 18). However, in the era of advanced HF therapy, there is increasing 5-year survival rates(19, 20). Currently, most of the literature on the incidence and prognosis of sarcoidosis patients with HF has been derived from single-center retrospective studies(21).

There remains a lack of large-population real-world data on the prognosis of hospitalized sarcoidosis patients with HF. Accordingly, we used the National Readmission Database (NRD) to identify the characteristics and clinical outcomes of sarcoidosis patients with heart failure (sw-HF) and after propensity matching, compared these to sarcoidosis patients without heart failure (sw/o-HF).

## METHODS

### Database selection

The NRD was developed by the Healthcare Cost and Utilization Project (HCUP) and sponsored by the Agency for Healthcare Research and Quality. The NRD data include inpatient care from all-payer hospitals and is the largest inpatient dataset in the United States of America (US)(22). It includes demographics, baseline characteristics including comorbidities, reasons for the admission, readmission days after the discharge, and reasons for the readmission. An advantage of utilizing these types of data is the ability to generate larger sample sizes for rarer diseases, for example CS, due to the large denominator as well as being representative of the US hospitalized population, reducing tertiary referral bias. This study was exempt from Institutional review board approval, given the use of deidentified, publicly available data.

### Population selection

Data on patients who were ≥18 years and admitted with sarcoidosis were abstracted from the NRD. Patients with presumed ischemic cardiomyopathy (ICM) were excluded from the data by removing those patients with a history of percutaneous coronary intervention (PCI) and/or coronary artery bypass grafting (CABG). The identification of admission for sarcoidosis (primary diagnosis) and the presence of HF from 2010 to 2019 were based on International Classification of Diseases-9th and 10th Revision-Clinical Modification (ICD-9-CM and ICD-10-CM, respectively; **Supplemental Table 1 and Figure 1**. Heart failure presence was defined as a documented history of either systolic or diastolic HF. Due to heterogenicity among admitting centers in coding for this diagnosis, there was no uniformity with regards to LVEF cut off in defining systolic HF or what imaging modality was applied during diagnosis.

**Figure 1.**
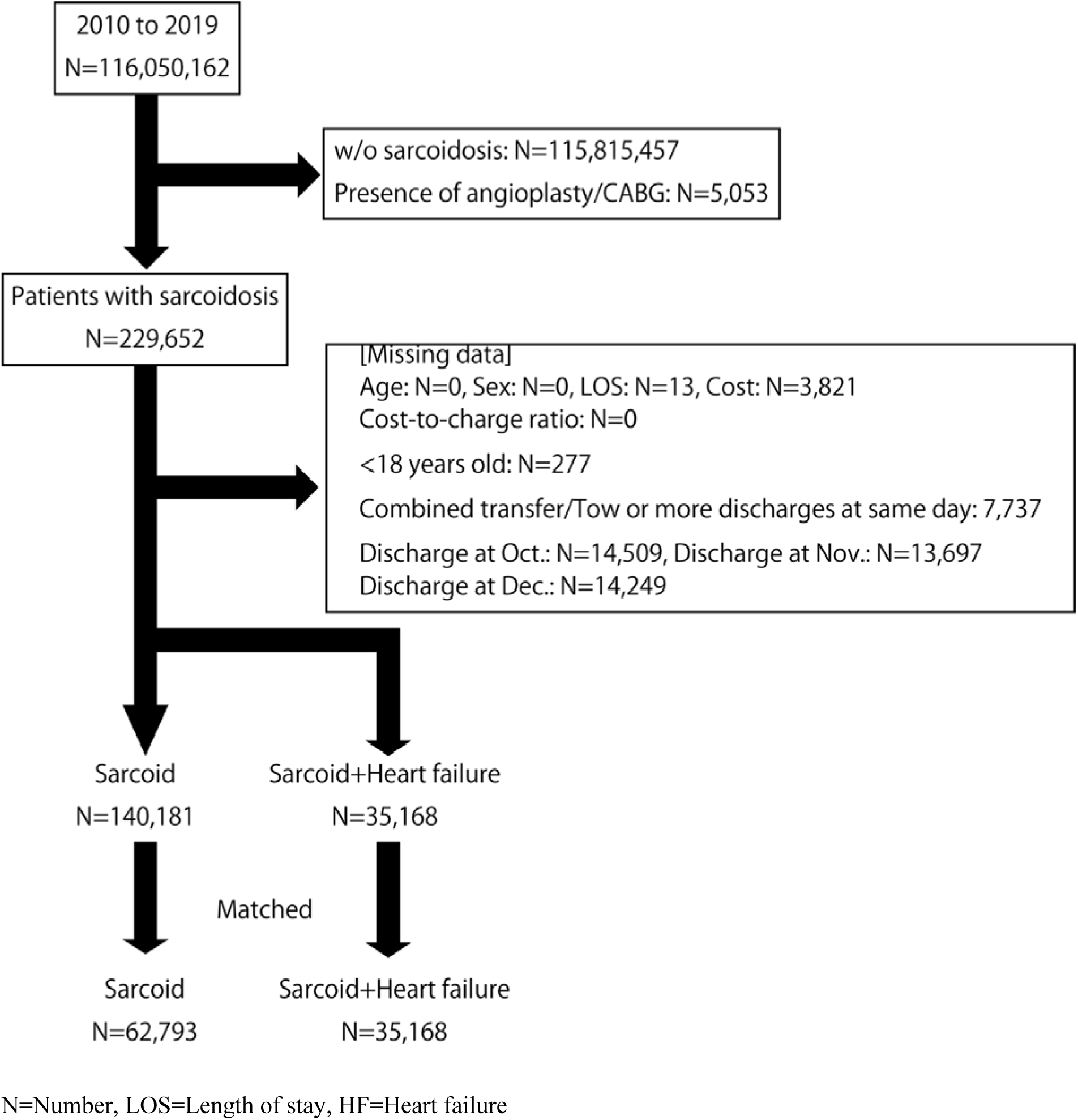
Inclusion Criteria Flow Chart

Initially, our study aimed to focus primarily on patients with ICD codes for CS. ICD-10 codes yielded very few CS patients from 2016-2019 (**Supplemental Table 2:** D8685 “sarcoid myocarditis”: 1252 hospitalizations). There were no ICD-9 codes specific for CS (for the years 2010-2015). Therefore, we changed the search strategy and after excluding patients with presumed ICM (ICD-10 codes 125.5 and 414), we identified all patients with extracardiac sarcoidosis plus HF. These patients were compared to matched cohort of sarcoidosis patients without HF.

Patients were followed up for 90-days from the point of discharge, with data limited to events within each calendar year. Those discharged from the months October to December were excluded due to 90-day follow-up not being available in these patients (**Figure 1**). Type of readmission (all-cause, atrial fibrillation [AF], ventricular tachycardia [VT], ventricular fibrillation [VF] and acute HF) were identified using the ICD-9-CM and ICD-10-CM codes (**Supplemental Table 3**). The presence of cardiac implanted electronic devices (CIED) was also recorded.

The primary payers were identified using the NRD database and categorized as Medicare/Medicaid, private insurance, and others. The medical costs of admission and readmission were calculated as adjusted medical costs, which were adjusted for the charge-to-cost ratio (provided by the HCUP) since the net medical costs included hospital bills for services such as wages, supplies, and utility that were different among the hospitals. The Charlson Comorbidities Index (CCI) was calculated using ICD-9-CM and ICD-10-CM codes (**Supplemental Table 5**)(23, 24).

### Statistical analysis

Descriptive statistics are presented as median (interquartile range; IQR) or number (%). Propensity score matching with the nearest neighbor method (1:2) and a maximum caliber of 0.20 of the score was used for adjusting for potential confounders (that is, adjusted for sex, age, and CCI) among the sw-HF and sw/o-HF. Comparisons between sw-HF and sw/o-HF were assessed using the χ2 test for categorical variables. Continuous data normality was tested using Shapiro–Wilk W test; and compared using Mann–Whitney U for non-normal. For estimation for freedom from readmission, the log-rank test and the Cox proportional hazards regression models were used. For competing risks analysis, we used Fine-Gray model (Figures 2B-D). Logistic regression analysis was used to estimate the risk for in-hospital mortality at second admission. As per NRD rules, to prevent identification, some cases were excluded if cases with n ≤10 in a particular hospital or area were present. The included variables for the Cox proportional hazards model and logistic regression analysis were age, sex, CCI, presence of comorbidities, and hospital bed size. Analysis was done using STATA v.15.1 (Stata-Corp). A two-tailed *a priori* p < 0.05 was regarded as significant.

**Figure 2.**
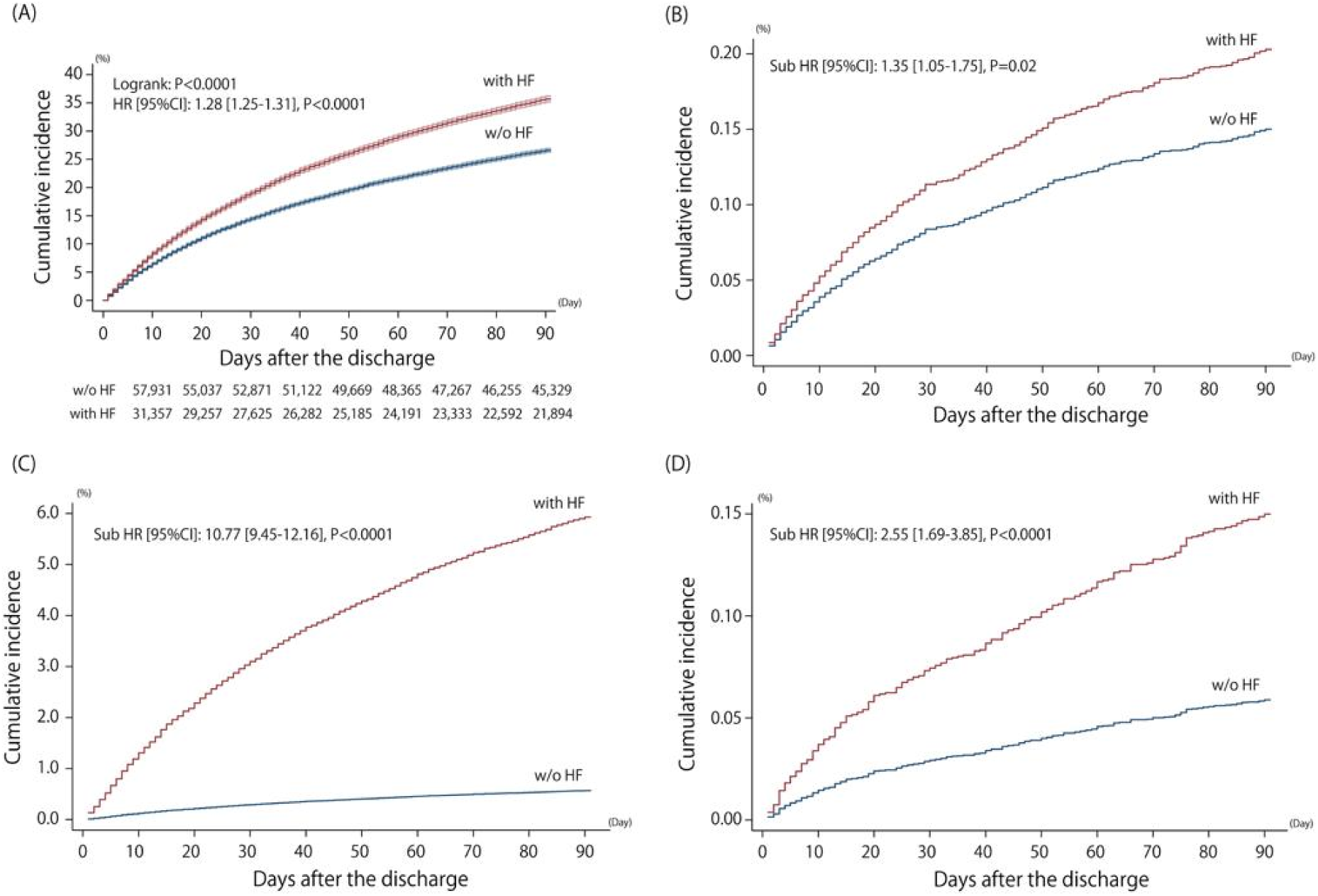
Kaplan-Meier Carve for 90 day readmission (B) Readmission of All events (C) Readmission of atrial fibrillation (D) Readmission of heart failure (E) Readmission of VT/VF HF=Intracardiac echocardiography HR=Hazard ratio, CI=Confidence Interval Shaded area indicates the 95% confidence interval. The hazard ratio and 95%CI were adjusted for age, sex, Charlson comorbidity index, CHA2DS2-VASc score and presence of anticoagulation therapy, comorbidities and hospital bed size.

## RESULTS

### Study population

Over the 10-year period (–2019), a total of 175,349 patients met the inclusion criterion out of whom 97,961 patients were included in the study after propensity matching and exclusion of cases that could have been identified (**Figure 1**).

### Baseline characteristics at time of index admission

The baseline characteristics of patients are summarized in **Table 1**. Out of the matched population, 35,168 (35.9%) sarcoidosis patients had a history of HF, whereas 62,793 (64.1%) patients did not. The median [IQR] age of the whole cohort was 63 [54-71] years with a Charlson Comorbidity Index of 3 [2-4] and a female preponderance (62.1%).

**Table 1.** Clinical Characteristics at 1st admission.

|  | All | w/o HF | with HF | P-value |
| --- | --- | --- | --- | --- |
| Number, N (%) | 97,961 (100.0) | 62,793 (64.1) | 35,168 (35.9) | - |
| Age, years | 63.0 (54.0-71.0) | 62.0 (53.0-71.0) | 64.0 (55.0-73.0) | <0.0001 |
| Male, N (%) | 37,097 (37.9) | 23,810 (37.9) | 13,287 (37.8) | 0.67 |
| Charlson comorbidity index, points | 3.0 (2.0-4.0) | 2.0 (2.0-3.0) | 3.0 (2.0-4.0) | <0.0001 |
| Anti-coagulant therapy, N (%) | 14,329 (14.6) | 6,775 (10.8) | 7,554 (21.5) | <0.0001 |
| Length of stay, day | 4.0 (2.0-6.0) | 4.0 (2.0-6.0) | 4.0 (3.0-7.0) | <0.0001 |
| **Healthcare associated cost, \$US** | | | | |
| Net value | 32,952.0<br>(17,962-62,581) | 31,594.0<br>(17,297.0-58,675.0) | 35,723.5<br>(19,329.0-70,509.0) | <0.0001 |
| Adjusted value | 9,232.2<br>(5,519.2-16,290.9) | 8,828.3<br>(5,303.1-15,384.9) | 9,954.5<br>(5,934.7-18,128.8) | <0.0001 |
| **CIED implantation, N (%)** |  |  |  |  |
| PPM | 539 (0.6) | 289 (0.5) | 250 (0.7) | <0.0001 |
| ICD | 629 (0.6) | 155 (0.3) | 474 (1.4) | <0.0001 |
| CRT-P | ≤50 (N/A) | ≤10 (N/A) | ≤40 (N/A) | <0.0001 |
| CRT-D | 301 (0.3) | 25 (0.04) | 276 (0.8) | <0.0001 |
| **Cardiac interventions, N (%)** |  |  |  |  |
| Catheter ablation | 372 (0.4) | 114 (0.2) | 258 (0.7) | <0.0001 |
| LVAD | ≤110 (N/A) | ≤10 (N/A) | ≤110 (N/A) | <0.0001 |
| Cardiac transplant | 24 (0.02) | 0 (0.0) | 24 (0.07) | <0.0001 |
| **Comorbidities, N (%)** |  |  |  |  |
| Ventricular tachycardia | 3,110 (3.2) | 820 (1.3) | 2,290 (6.5) | <0.0001 |
| Ventricular fibrillation | 247 (0.3) | 91 (0.1) | 156 (0.4) | <0.0001 |
| Atrioventricular block | 2,043 (2.1) | 883 (1.4) | 1,160 (3.3) | <0.0001 |
| Atrial fibrillation | 12,464 (12.7) | 4,703 (7.5) | 7,761 (22.1) | <0.0001 |
| Cerebrovascular diseases | 6,309 (6.4) | 4,721 (7.5) | 1,588 (4.5) | <0.0001 |
| Chronic Pulmonary disease | 46,501 (47.5) | 30,136 (48.0) | 16,365 (46.5) | <0.0001 |
| Peripheral vascular diseases | 8,165 (8.3) | 3,931 (6.3) | 4,234 (12.0) | <0.0001 |
| Liver diseases | 6,878 (7.0) | 5,205 (8.3) | 1,673 (4.8) | <0.0001 |
| Diabetes | 43,515 (44.4) | 29,241 (46.6) | 14,274 (40.6) | <0.0001 |
| Renal diseases | 31,545 (32.2) | 18,677 (29.7) | 12,868 (36.6) | <0.0001 |
| Hypertension | 61,106 (62.4) | 45,170 (71.9) | 15,936 (45.3) | <0.0001 |
| Coagulation defects | 1,104 (1.1) | 680 (1.1) | 424 (1.2) | 0.08 |
| Obesity | 22,800 (23.3) | 13,091 (20.9) | 9,709 (27.6) | <0.0001 |
| **Hospital bed size, N (%)** |  |  |  |  |
| Small | 12,805 (13.1) | 8,272 (13.2) | 4,533 (12.9) |  |
| Medium | 26,403 (27.0) | 17,020 (27.1) | 9,383 (26.7) |  |
| Large | 58,753 (60.0) | 37,501 (59.7) | 21,252 (60.4) | 0.09 |
| **Primary payer, N (%)** |  |  |  |  |
| Medicare/Medicaid | 69,987 (71.6) | 43,071 (68.7) | 26,916 (76.7) |  |
| Private insurance | 22,646 (23.2) | 16,153 (25.8) | 6,493 (18.5) |  |
| Others | 5,152 (5.3) | 3,459 (5.5) | 1,693 (4.8) | <0.0001 |

HF=Intracardiac echocardiography, N=Number, HAC=Healthcare associated cost

HF patients had a significantly higher prevalence of AVB (3.3 vs. 1.4%, p <0.0001), previous VT (6.5 vs. 1.3%, p <0.0001), previous VF (0.4 vs. 0.1%, p <0.0001) and AF (22.1 vs 7.5%, p <0.0001). They had a similar, though statistically greater, median length of stay (LOS) (4 [3-7] vs. 4 [2-6] days, p <0.0001) and higher median adjusted healthcare-associated costs (HAC) (US$ 9,954.5 [5,934.7-18,128.8] vs. 8,828.3 [5,303.1-15,384.9], p <0.0001) in the index admission when compared to those without HF.

On first admission, CIED were more common in HF patients (permanent pacemaker [PPM] 0.7 vs 0.5%, implantable cardioverter defibrillator 1.4 vs 0.3%, and Cardiac Resynchronization Therapy-Defibrillators (CRT-D) 0.8 vs 0.04%, all p<0.0001). Moreover, a higher number of HF patients underwent catheter ablation (0.7 vs. 0.2%, p <0.0001) or cardiac transplantation (0.07 vs 0.0%, p<0.0001) during the index admission. Peripheral vascular disease, renal impairment, obesity and anticoagulation use were also more prevalent in patients with HF (p <0.0001). Conversely, cerebrovascular disease, chronic pulmonary disease, liver disease, diabetes and hypertension were more commonly observed in those without HF (p <0.0001). With regards to payment plans, Medicare/Medicaid insurance was more common (76.7 vs. 68.7%, p<0.0001) among HF patients whereas private insurance use was less frequent (18.5 vs. 25.8%, p <0.0001).

### Comparison between patients with and without 90-day readmission

Across both HF and non-HF sarcoidosis patients, those who were readmitted within 90-days had higher Charlson Comorbidity Index, longer LOS, higher HAC and greater use of anticoagulant therapy on index admission **(Table 2).** HF patients had a higher proportion of index admissions with VT/VF (1.5 vs. 0.3%, p <0.0001), AVB (0.4 vs 0.3%, p <0.0001, AF (1.9 vs. 0.8%, p <0.0001) and cardiac arrest (0.06% vs 0.02%, p=0.003) compared to those without HF (**Table 3A**). There was a greater proportion of HF patients undergoing 2^nd^ admission within 90 days for, VT/VF, AF or acute HF (all p<0.0001) compared to non-HF patients. (**Table 3B**).

**Table 2.**
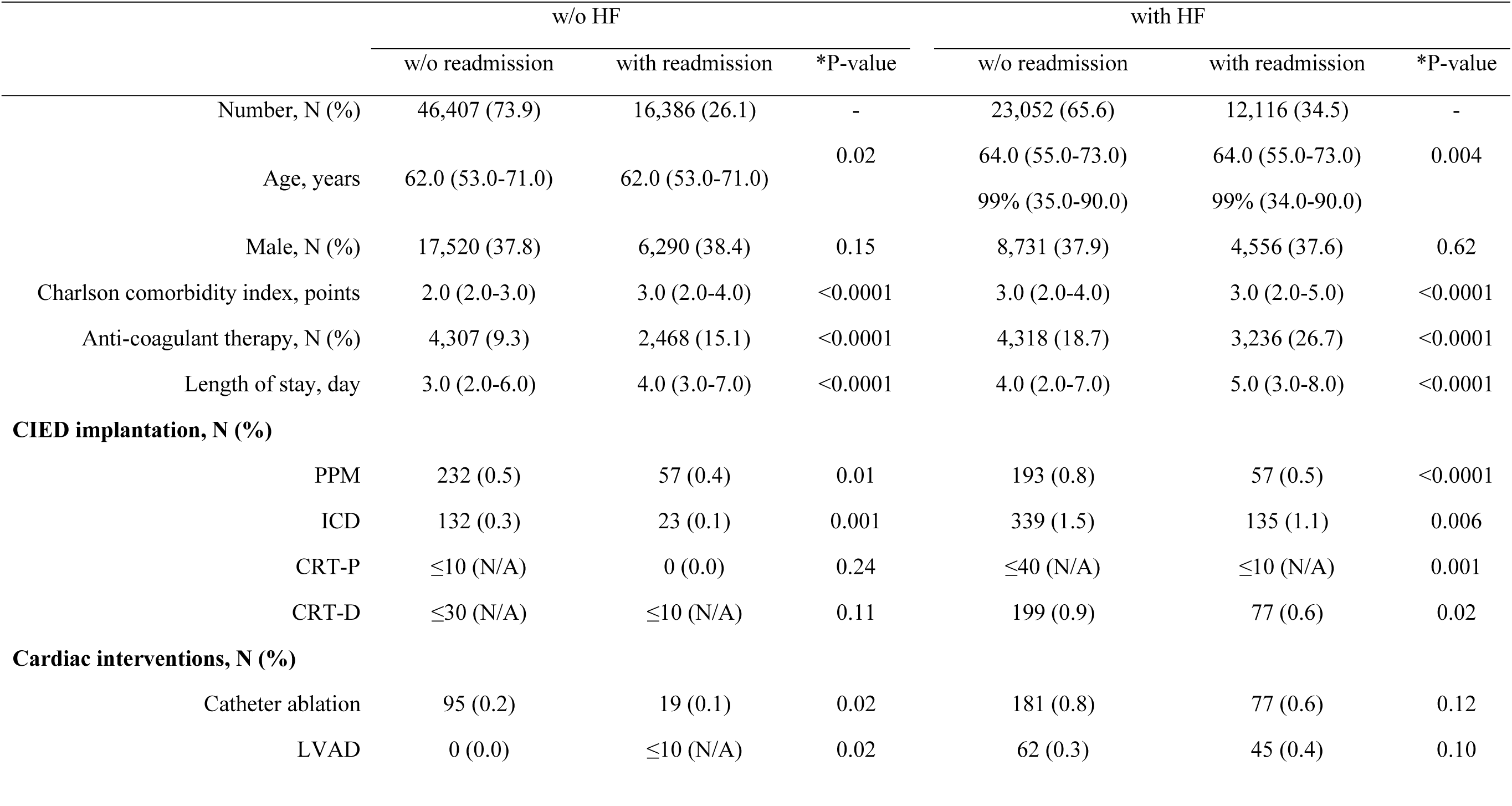
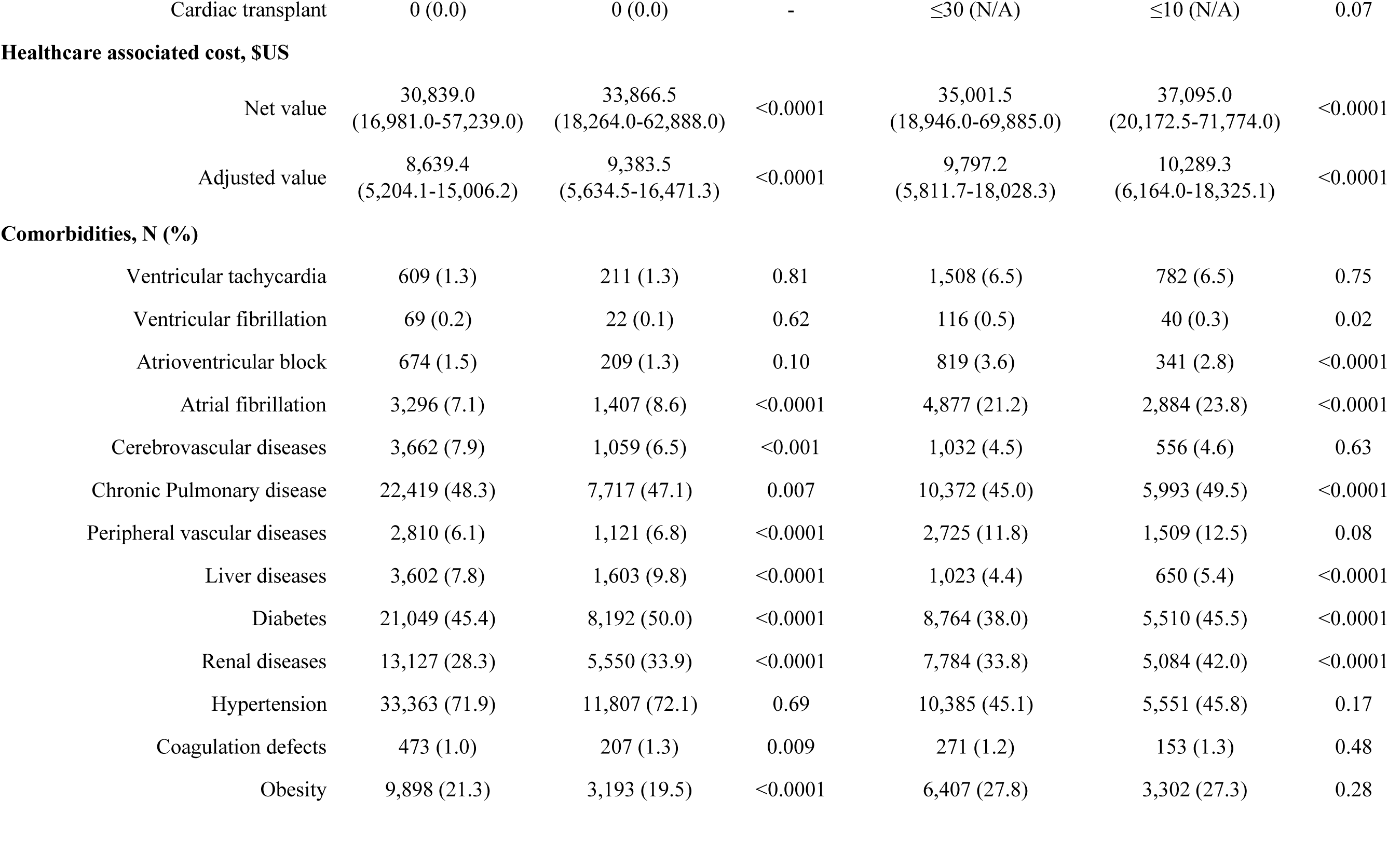
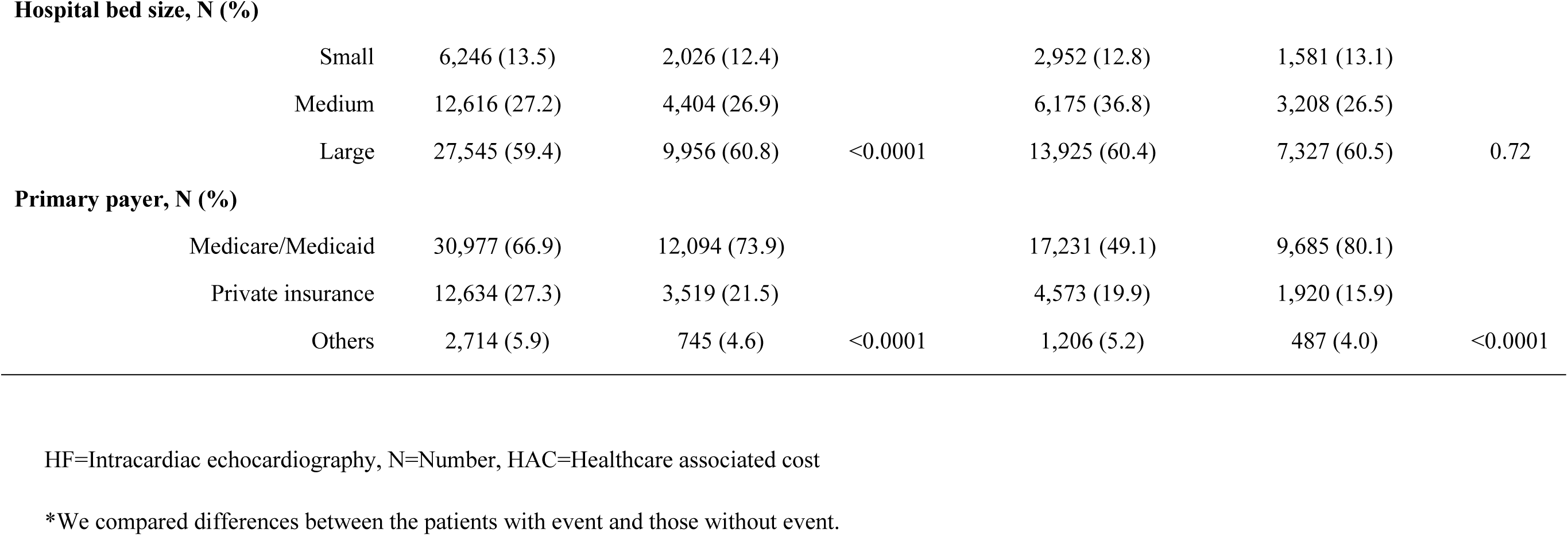
Differences on index admission observed between patients with readmissions and those without readmissions at 90 days.

|  | w/o HF |  |  | with HF |  |  |
| --- | --- | --- | --- | --- | --- | --- |
|  | w/o readmission | with readmission | *P-value | w/o readmission | with readmission | *P-value |
| Number, N (%) | 46,407 (73.9) | 16,386 (26.1) | - | 23,052 (65.6) | 12,116 (34.5) | - |
| Age, years | 62.0 (53.0-71.0) | 62.0 (53.0-71.0) | 0.02 | 64.0 (55.0-73.0) | 64.0 (55.0-73.0) | 0.004 |
|  |  |  |  | 99% (35.0-90.0) | 99% (34.0-90.0) |  |
| Male, N (%) | 17,520 (37.8) | 6,290 (38.4) | 0.15 | 8,731 (37.9) | 4,556 (37.6) | 0.62 |
| Charlson comorbidity index, points | 2.0 (2.0-3.0) | 3.0 (2.0-4.0) | <0.0001 | 3.0 (2.0-4.0) | 3.0 (2.0-5.0) | <0.0001 |
| Anti-coagulant therapy, N (%) | 4,307 (9.3) | 2,468 (15.1) | <0.0001 | 4,318 (18.7) | 3,236 (26.7) | <0.0001 |
| Length of stay, day | 3.0 (2.0-6.0) | 4.0 (3.0-7.0) | <0.0001 | 4.0 (2.0-7.0) | 5.0 (3.0-8.0) | <0.0001 |
| **CIED implantation, N (%)** |  |  |  |  |  |  |
| PPM | 232 (0.5) | 57 (0.4) | 0.01 | 193 (0.8) | 57 (0.5) | <0.0001 |
| ICD | 132 (0.3) | 23 (0.1) | 0.001 | 339 (1.5) | 135 (1.1) | 0.006 |
| CRT-P | ≤10 (N/A) | 0 (0.0) | 0.24 | ≤40 (N/A) | ≤10 (N/A) | 0.001 |
| CRT-D | ≤30 (N/A) | ≤10 (N/A) | 0.11 | 199 (0.9) | 77 (0.6) | 0.02 |
| **Cardiac interventions, N (%)** |  |  |  |  |  |  |
| Catheter ablation | 95 (0.2) | 19 (0.1) | 0.02 | 181 (0.8) | 77 (0.6) | 0.12 |
| LVAD | 0 (0.0) | ≤10 (N/A) | 0.02 | 62 (0.3) | 45 (0.4) | 0.10 |
| Cardiac transplant | 0 (0.0) | 0 (0.0) | - | ≤30 (N/A) | ≤10 (N/A) | 0.07 |
| **Healthcare associated cost, \$US** | | | | | | |
| Net value | 30,839.0<br>(16,981.0-57,239.0) | 33,866.5<br>(18,264.0-62,888.0) | <0.0001 | 35,001.5<br>(18,946.0-69,885.0) | 37,095.0<br>(20,172.5-71,774.0) | <0.0001 |
| Adjusted value | 8,639.4<br>(5,204.1-15,006.2) | 9,383.5<br>(5,634.5-16,471.3) | <0.0001 | 9,797.2<br>(5,811.7-18,028.3) | 10,289.3<br>(6,164.0-18,325.1) | <0.0001 |
| **Comorbidities, N (%)** |  |  |  |  |  |  |
| Ventricular tachycardia | 609 (1.3) | 211 (1.3) | 0.81 | 1,508 (6.5) | 782 (6.5) | 0.75 |
| Ventricular fibrillation | 69 (0.2) | 22 (0.1) | 0.62 | 116 (0.5) | 40 (0.3) | 0.02 |
| Atrioventricular block | 674 (1.5) | 209 (1.3) | 0.10 | 819 (3.6) | 341 (2.8) | <0.0001 |
| Atrial fibrillation | 3,296 (7.1) | 1,407 (8.6) | <0.0001 | 4,877 (21.2) | 2,884 (23.8) | <0.0001 |
| Cerebrovascular diseases | 3,662 (7.9) | 1,059 (6.5) | <0.001 | 1,032 (4.5) | 556 (4.6) | 0.63 |
| Chronic Pulmonary disease | 22,419 (48.3) | 7,717 (47.1) | 0.007 | 10,372 (45.0) | 5,993 (49.5) | <0.0001 |
| Peripheral vascular diseases | 2,810 (6.1) | 1,121 (6.8) | <0.0001 | 2,725 (11.8) | 1,509 (12.5) | 0.08 |
| Liver diseases | 3,602 (7.8) | 1,603 (9.8) | <0.0001 | 1,023 (4.4) | 650 (5.4) | <0.0001 |
| Diabetes | 21,049 (45.4) | 8,192 (50.0) | <0.0001 | 8,764 (38.0) | 5,510 (45.5) | <0.0001 |
| Renal diseases | 13,127 (28.3) | 5,550 (33.9) | <0.0001 | 7,784 (33.8) | 5,084 (42.0) | <0.0001 |
| Hypertension | 33,363 (71.9) | 11,807 (72.1) | 0.69 | 10,385 (45.1) | 5,551 (45.8) | 0.17 |
| Coagulation defects | 473 (1.0) | 207 (1.3) | 0.009 | 271 (1.2) | 153 (1.3) | 0.48 |
| Obesity | 9,898 (21.3) | 3,193 (19.5) | <0.0001 | 6,407 (27.8) | 3,302 (27.3) | 0.28 |
| **Hospital bed size, N (%)** |  |  |  |  |  |  |
| Small | 6,246 (13.5) | 2,026 (12.4) |  | 2,952 (12.8) | 1,581 (13.1) |  |
| Medium | 12,616 (27.2) | 4,404 (26.9) |  | 6,175 (36.8) | 3,208 (26.5) |  |
| Large | 27,545 (59.4) | 9,956 (60.8) | <0.0001 | 13,925 (60.4) | 7,327 (60.5) | 0.72 |
| **Primary payer, N (%)** |  |  |  |  |  |  |
| Medicare/Medicaid | 30,977 (66.9) | 12,094 (73.9) |  | 17,231 (49.1) | 9,685 (80.1) |  |
| Private insurance | 12,634 (27.3) | 3,519 (21.5) |  | 4,573 (19.9) | 1,920 (15.9) |  |
| Others | 2,714 (5.9) | 745 (4.6) | <0.0001 | 1,206 (5.2) | 487 (4.0) | <0.0001 |

HF=Intracardiac echocardiography, N=Number, HAC=Healthcare associated cost
\*We compared differences between the patients with event and those without event.

**Table 3.** Reasons for admission.

**(A) First admission**
|  | w/o HF | with HF | P-value |
| --- | --- | --- | --- |
| VT/VF | 208 (0.3) | 527 (1.5) | <0.0001 |
| Atrioventricular block | 168 (0.3) | 150 (0.4) | <0.0001 |
| Atrial fibrillation | 494 (0.8) | 653 (1.9) | <0.0001 |
| Acute Heart Failure | 0 (0.0) | 7,558 (21.5) | <0.0001 |
| Cardiac arrest | 14 (0.02) | 21 (0.06) | 0.003 |

**(B) Second admission**
|  | w/o HF | with HF | P-value |
| --- | --- | --- | --- |
| VT/VF | 52 (0.3) | 183 (1.5) | <0.0001 |
| Atrioventricular block | ≤10 (N/A) | 13 (0.1) | 0.17 |
| Atrial fibrillation | 140 (0.9) | 213 (1.8) | <0.0001 |
| Acute Heart Failure | 367 (2.2) | 2,307 (19.0) | <0.0001 |
| Cardiac arrest | ≤10 (N/A) | ≤10 (N/A) | 0.41 |

Values are shown as number (%).

### Predictors of 90-day readmission

On univariable Cox proportional hazard modelling, history of AVB (HR 0.87 [0.80-0.94], p=0.001) and the presence of an existing CIED (HR 0.70 [0.57-0.85], p=0.001) on index admission were associated with freedom from 90-days readmission. Increasing age, higher Charlson Comorbidity Index and use of anticoagulant therapy were predictive of readmission. Additionally, the presence of AF, chronic pulmonary disease, liver disease, diabetes, renal disease and coagulation defects were also predictors of 90-day readmission (**Table 4**).

**Table 4.**
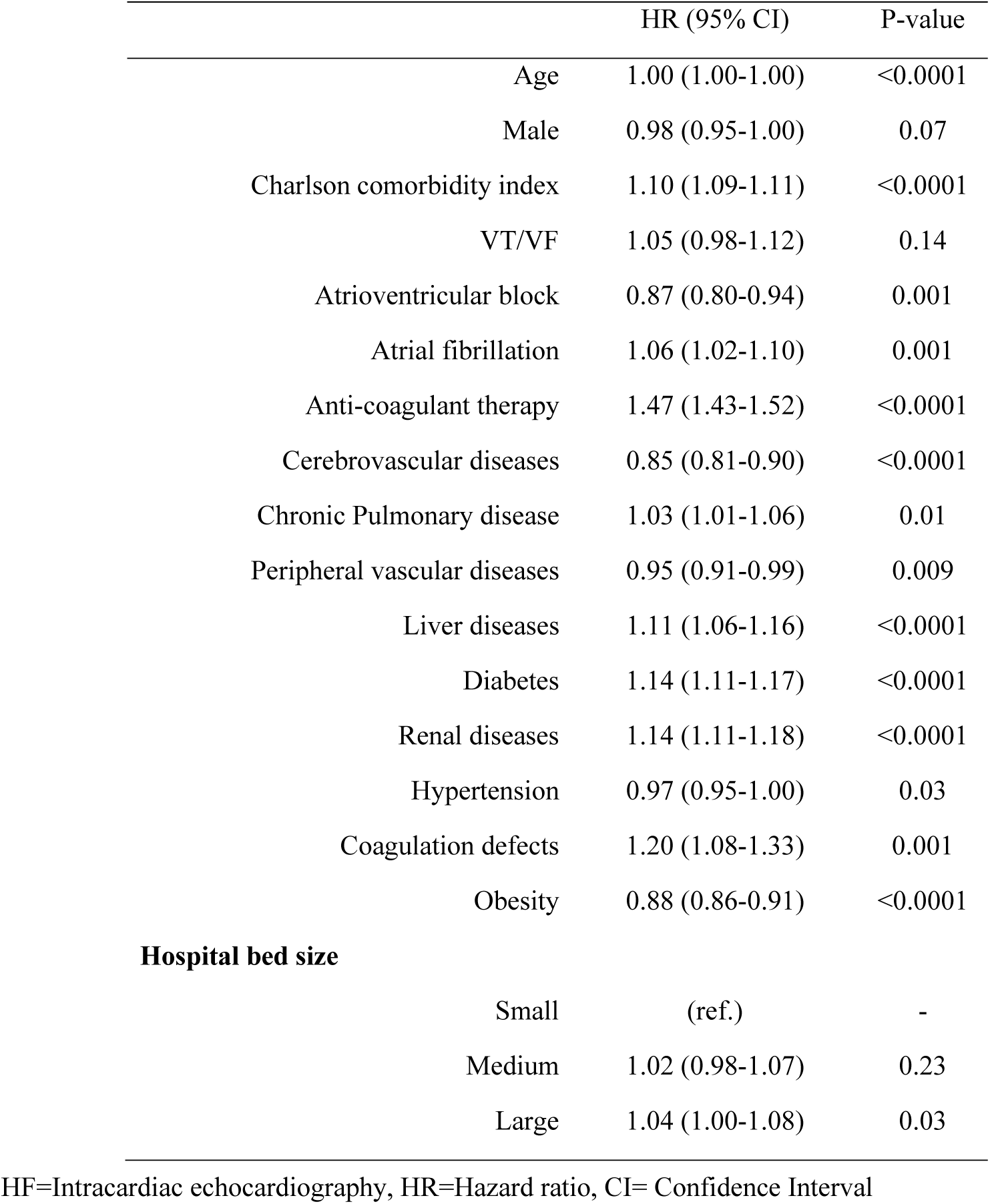
Predictors at 1st admission for readmission.

The Kaplan-Meier curves in **Figure 2** demonstrate that HF patients had higher incidence of readmissions within 90-days when compared to non-HF patients. This was consistent for all-cause readmissions (HR 1.28 [1.25–1.31], p< 0.0001) as well as readmissions due to AF (HR 1.35 [1.05-1.75], p=0.02), acute HF (HR 10.77 [9.45–12.16], p< 0.0001) or VT/VF (2.55 [1.69–3.85], p <0.0001) **(Figure 2A-D).**

### Mortality, length of stay and cost of readmission

Rates of in-hospital mortality on readmission was higher among HF patients (5.1 vs. 3.8%, p < 0.0001). On univariable analysis, the predictors of in-hospital mortality at readmission were HF presence (OR 1.24 [1.09-1.41], p=0.001), increasing age (OR 1.03 [1.02-1.03], p < 0.0001), male sex (OR 1.20 [1.06-1.35], p=0.003), higher Charlson Comorbidity Index (OR 1.14 [1.10-1.17], p < 0.0001) and presence of liver disease (OR 1.29 [1.06-1.57], p=0.01), (**Tables 5 and 6**). Additionally, the median LOS (5 [3-8] vs. 4 [3-7] days, p< 0.0001) and HAC (US$ 9,667.0 [5,626.6-18,076.6] vs. 9,087.1 [5,378.3-16,966.1], p < 0.0001) upon readmission were significantly higher in HF patients (**Table 5**).

**Table 5.**
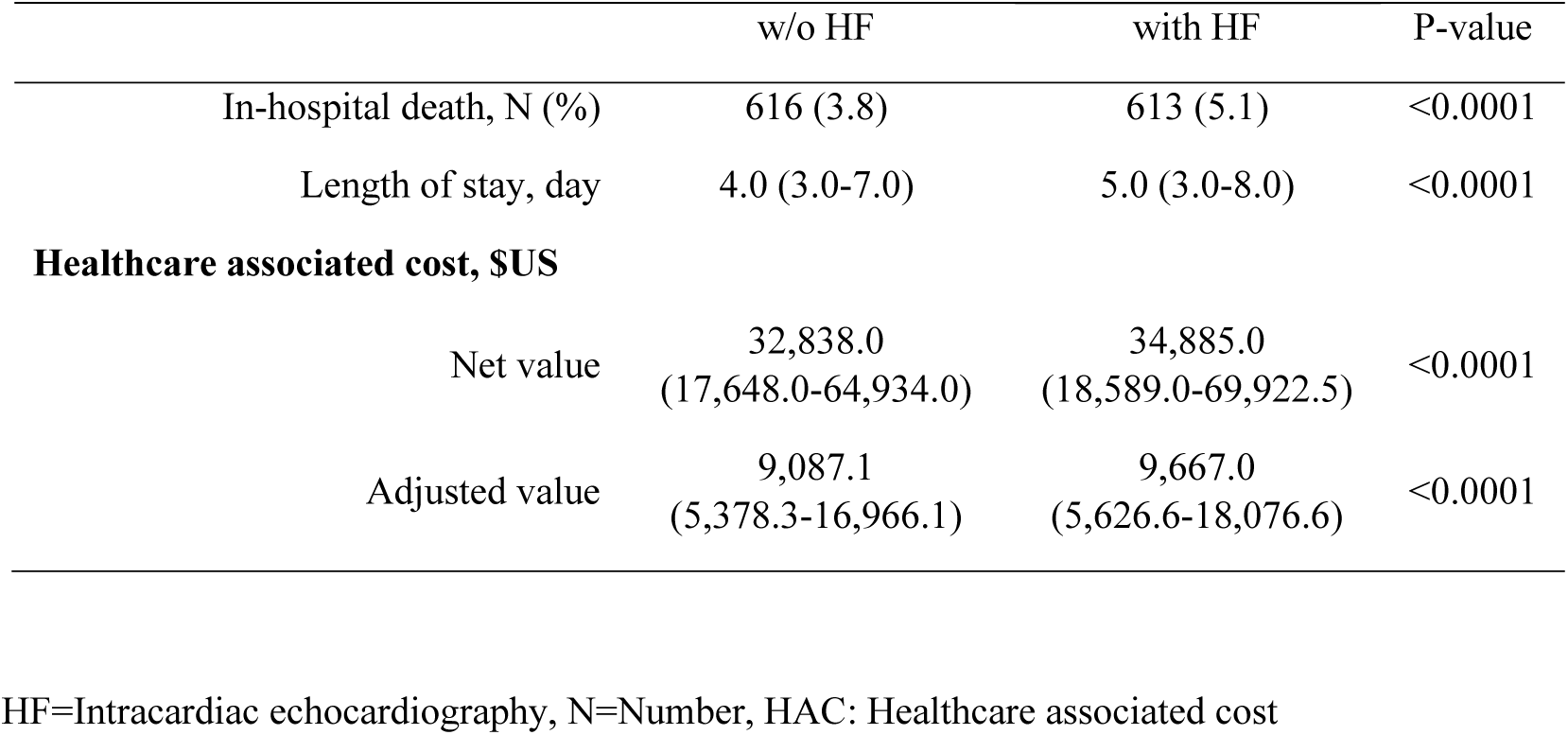
Differences between patients with HF and those without HF at readmission.

|  | w/o HF | with HF | P-value |
| --- | --- | --- | --- |
| In-hospital death, N (%) | 616 (3.8) | 613 (5.1) | <0.0001 |
| Length of stay, day | 4.0 (3.0-7.0) | 5.0 (3.0-8.0) | <0.0001 |
| **Healthcare associated cost, \$US** | | | |
| Net value | 32,838.0<br>(17,648.0-64,934.0) | 34,885.0<br>(18,589.0-69,922.5) | <0.0001 |
| Adjusted value | 9,087.1<br>(5,378.3-16,966.1) | 9,667.0<br>(5,626.6-18,076.6) | <0.0001 |

HF=Intracardiac echocardiography, N=Number, HAC: Healthcare associated cost

**Table 6.** Predictors of increasing LOS, HAC, and in-hospital death at readmission.

|  | LOS |  | Adjusted HAC |  | In-hospital death |  |
| --- | --- | --- | --- | --- | --- | --- |
| | $\beta$ | P-value | $\beta$ | P-value | OR (95%CI) | P-value |
| HF | 0.015 | 0.02 | 0.014 | 0.04 | 1.24 (1.09-1.41) | 0.001 |
| Age | 0.009 | 0.14 | 0.002 | 0.71 | 1.03 (1.02-1.03) | <0.0001 |
| Male | 0.001 | 0.86 | 0.018 | 0.004 | 1.20 (1.06-1.35) | 0.003 |
| Charlson comorbidity index | 0.023 | 0.002 | 0.017 | 0.02 | 1.14 (1.10-1.17) | <0.0001 |
| Anti-coagulant therapy | -0.006 | 0.35 | 0.006 | 0.31 | 0.76 (0.65-0.89) | 0.001 |
| VT/VF | 0.006 | 0.33 | 0.036 | <0.0001 | 1.02 (0.75-1.38) | 0.90 |
| Atrioventricular block | 0.027 | 0.65 | 0.020 | 0.001 | 0.81 (0.52-1.26) | 0.35 |
| Atrial fibrillation | 0.009 | 0.14 | 0.004 | 0.51 | 1.09 (0.92-1.27) | 0.32 |
| Cerebrovascular diseases | 0.012 | 0.048 | 0.0003 | 0.96 | 0.79 (0.61-1.03) | 0.08 |
| Chronic Pulmonary disease | -0.034 | <0.0001 | -0.042 | <0.0001 | 0.91 (0.81-1.02) | 0.11 |
| Peripheral vascular diseases | -0.003 | 0.66 | 0.020 | 0.001 | 0.76 (0.62-0.94) | 0.01 |
| Liver diseases | 0.010 | 0.11 | 0.022 | <0.0001 | 1.29 (1.06-1.57) | 0.01 |
| Diabetes | 0.001 | 0.85 | -0.017 | 0.006 | 0.75 (0.66-0.84) | <0.0001 |
| Renal diseases | 0.019 | 0.007 | 0.008 | 0.27 | 0.87 (0.77-0.99) | 0.04 |
| Hypertension | -0.018 | 0.004 | -0.035 | <0.0001 | 0.86 (0.76-0.97) | 0.02 |
| Coagulation defects | 0.023 | <0.0001 | 0.018 | 0.002 | 1.40 (0.91-2.16) | 0.13 |
| Obesity | 0.007 | 0.22 | 0.007 | 0.26 | 0.75 (0.64-0.88) | <0.0001 |
| **Hospital bed size** |  |  |  |  |  |  |
| Small | (ref.) | - | (ref.) | - | (ref.) | - |
| Medium | -0.001 | 0.91 | -0.011 | 0.21 | 1.12 (0.92-1.37) | 0.27 |
| Large | 0.042 | <0.0001 | 0.038 | <0.0001 | 1.17 (0.97-1.40) | 0.11 |

## DISCUSSION

This is the largest study of an inpatient cohort of propensity-matched sarcoidosis patients with and without HF and has several important findings. First, HF in patients with sarcoidosis was associated with a significantly greater prevalence of conduction disease, ventricular arrhythmias, atrial fibrillation and presence of CIEDs on index presentation. Second, the healthcare adjusted costs were significantly greater in sarcoidosis patients with HF compared to those without, both on index admission and on 90-day readmission. Third, at the time of readmission, LOS and in-patient mortality was greater among HF patients. Fourth, HF patients were more likely to be readmitted within 90 days for all causes, including acute heart failure, AF and VT/VF. Lastly, among HF patients with known AVB, a 90-day readmission was less likely than those without AVB, likely due to those with conduction disease receiving a CIED during the index admission.

### Sarcoidosis and heart failure

HF has an incidence of 240 per 1000 patient-years in sarcoidosis and 12 per 1000 patient-years in patients without sarcoidosis(25). The etiology is multifactorial and can be with reduced or preserved ejection fraction(26, 27). Symptomatic HF in these patients provokes a decline in functional capacity and quality of life which are important reasons for hospitalization(28). While our study lacked the ability to focus solely on confirmed CS cases, there is likely to be a significant proportion of the sarcoidosis plus HF patients who do indeed have CS. CS patients with or without LV systolic dysfunction are at greater risk of ventricular arrhythmias and high-grade AVB. However, those with LVEF <40% display the lowest survival rates(29). It is estimated that HF accounts for nearly a quarter of deaths in CS(30). We found that in sarcoidosis patients, HF presence was linked to increased prevalence of arrhythmias, conduction disorders and atrial fibrillation. This in turn translated to increased LOS, greater healthcare costs and increased mortality. Our results highlight the additional burden placed on the patient as well as healthcare systems with two major comorbidities. One of the cornerstones of sarcoidosis management is immunosuppressive therapy. However, the additional fluid retention associated with corticosteroids can be deleterious in HF. Furthermore, 2^nd^ and 3^rd^ line treatments such as Tumour Necrosis Factor alpha (TNF-a) inhibitors can be associated with worsening LVSD in rare cases(31). Therefore, not only does the combination of sarcoidosis and HF encompass a group of CS patients at risk of arrhythmic SCD, the sarcoidosis treatments required can occasionally exacerbate acute HF episodes. While we compared sarcoidosis patients with and without HF, other investigators have examined hospital episodes for sarcoidosis patients with and without arrhythmias. *Desai et al* identified that those with arrhythmias had significantly higher mortality (3.7 vs. 1.5%), greater hospital LOS (6.4 vs. 5.2 days), and larger hospital charges ($64,118 *vs.* $41,565) in comparison to patients without arrhythmias (p < 0.001)(32). This emphasizes the need for robust anti-arrhythmic surveillance and management in patients with sarcoidosis.

### Corticosteroids to improve outcomes in cardiac sarcoidosis

A study conducted on 95 Japanese patients with CS demonstrated that HF severity was an important independent predictor of mortality with a 77% rise in death per increase in NYHA class. In this study, immunosuppression with corticosteroid therapy prior to the onset of systolic dysfunction led to improved clinical outcomes(33). *Kato et al* reported on 40 CS patients of whom half presented with high-grade AVB with preserved LV systolic function. Those treated with corticosteroids demonstrated improved survival(34). Similar findings were demonstrated in another Japanese study carried out on CS patients out of whom 31 experienced premature ventricular complexes (PVCs) and 14 participants suffered from non-sustained VT (NSVT). Participants having LVEF>35% demonstrated a significant reduction in PVCs and NSVT after treatment with prednisolone(35). All these previous studies were retrospective small, single-center studies. Our study adds to the literature using a large real world readmissions database that HF in patients with sarcoidosis have worse outcomes.

### Clinical indicators of inpatient mortality in Sarcoidosis

Our findings demonstrate that HF, advancing age, male gender, higher Charlson Comorbidity Index and the presence of liver disease are predictors of in-patient mortality once a sarcoidosis patient is readmitted to hospital. Prior studies have reported increasing age to be an independent risk factor for inpatient mortality among sarcoidosis patients(36). Similar to our findings, female gender appeared to be a protective factor. A key demographic not examined in our study was ethnicity, due to limitations within the NRD database(37). However, across the US, Black patients are known to have poorer health outcomes compared to Caucasians, particularly among those with sarcoidosis, where Black ethnicity was linked to 21% increase in in-hospital mortality(36).

### Strengths and limitations

The major strengths of our study are the large number of patients who were not confined to a single specialist center. Instead, this is a nationally representative sample thus reducing tertiary referral bias. Another strength was the robust data obtained regarding HAC and LOS and hospital readmissions which allowed us to assess the impact of clinical variables on the healthcare system. By excluding patients with ischemic heart disease, we increased the chances that the majority of patients captured with heart failure and sarcoidosis had suspected CS. However, in the absence of histological confirmation, compatible late gadolinium enhancement on cardiac magnetic resonance imaging (CMR) and suggestive uptake on ^18^F-fludeoxyglucose positron emission tomography (FDG-PET), no CS diagnostic criteria could be met.

Therefore, other overlapping diseases such as arrhythmogenic or LAMIN cardiomyopathy could not be excluded. Our study had several other limitations. We could not exclude misclassification bias which are inherent to such large databases. A key issue was the heterogenicity in labelling HF, as this included both systolic and diastolic presentations. Furthermore, the diagnosis and coding of sarcoidosis, particularly CS has been historically poor. While CS is mostly diagnosed using advanced imaging modalities such as CMR and FDG-PET, this information was not available from the NRD database and as such ICD codes were used. Information on the use of HF and immunosuppression therapies by sarcoidosis patients was unavailable and therefore the impact of such treatment on patient outcomes could not be evaluated.

### Future Directions

Given that we have demonstrated significantly worse mortality, cost and length of stay in sarcoidosis patients with heart failure, our study calls for screening for cardiac involvement in those patients with systemic sarcoidosis or *de novo* presentation with HF in patients suspected to have isolated CS. This can be done using screening patient symptoms, 12-lead electrocardiogram, brain natriuretic peptide (BNP) levels and echocardiography(38, 39). In order to improve outcomes, we propose the early implementation of prognostically beneficial HF medications and device therapies where relevant, in this patient cohort.

## Conclusion

Heart failure in hospitalized sarcoidosis patients is associated with increased prevalence of arrythmia, conduction disorders and a significantly higher length of stay, healthcare-adjusted costs, 90-days readmissions and mortality following readmission. Prospective studies are necessary to validate these findings and develop preventative strategies in order to reduce mortality and healthcare burdens.

## Data Availability

All manuscript authors confirm the following: 1) The paper is not under consideration elsewhere 2) None of the paper's contents have been previously published 3) All authors have read and approved the manuscript 4) All potential conflicts of interest have been disclosed.

## ACKNOWLEDGMENT

This research did not receive any specific grant from funding agencies in the public, commercial, or not-for-profit sectors.

## CENTRAL ILLUSTRATION

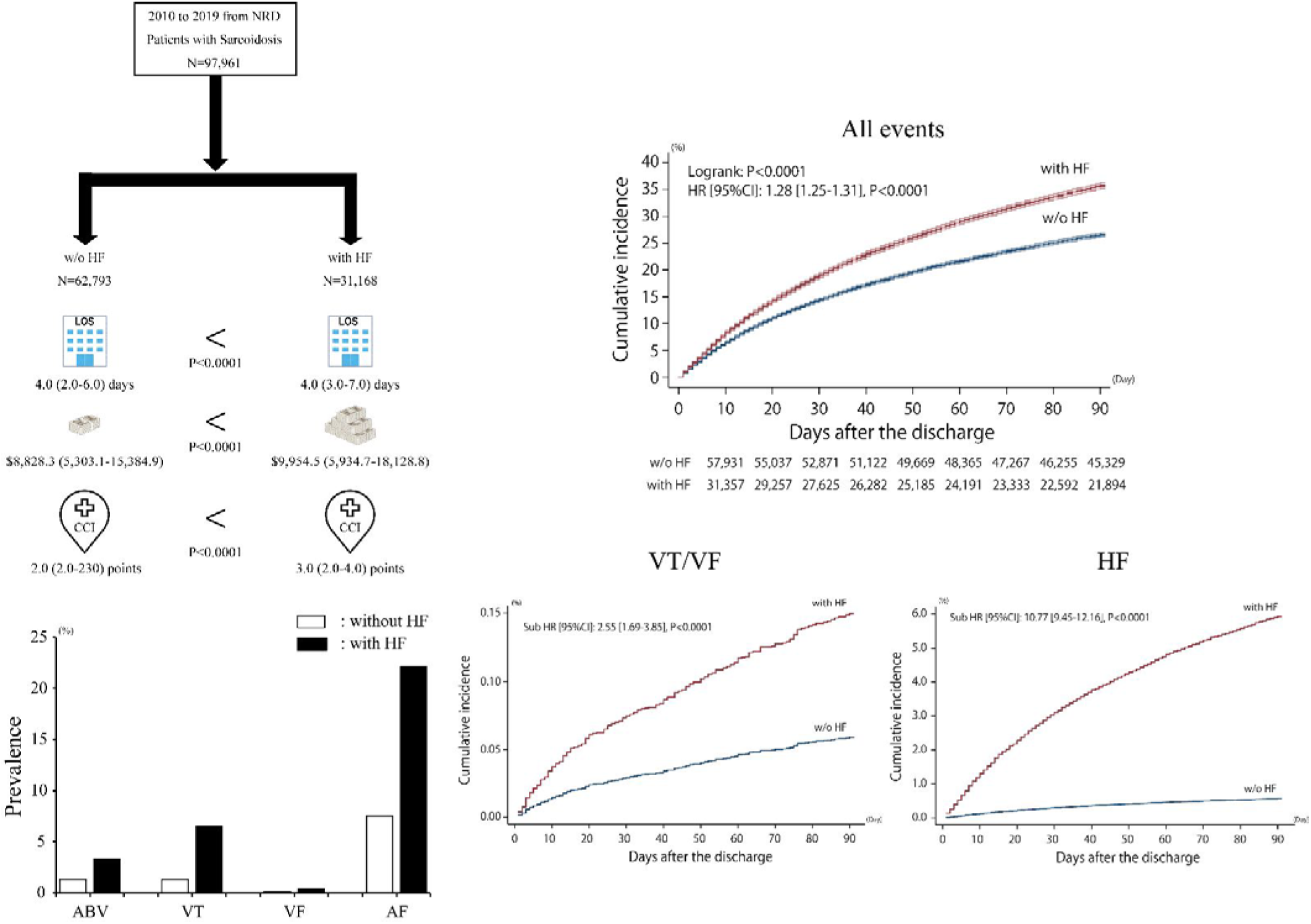

## References

1. Sikjær MG, Hilberg O, Ibsen R, Løkke A. Sarcoidosis: A nationwide registry-based study of incidence, prevalence and diagnostic work-up. Respir Med. 2021;187:106548.

2. Arkema EV, Cozier YC. Epidemiology of sarcoidosis: current findings and future directions. Ther Adv Chronic Dis. 2018;9(11):227–40.

3. Hunninghake G, Costabel U, Ando M, Baughman R, Cordier J, duBois R, et al. ATS/ERS/WASOG statement on sarcoidosis. American Thoracic Society/European Respiratory Society/World Association of Sarcoidosis and other Granulomatous Disorders. Sarcoidosis Vasc Diffuse Lung Dis. 1999;16:149–73.

4. Perry A, Vuitch F. Causes of death in patients with sarcoidosis. A morphologic study of 38 autopsies with clinicopathologic correlations. Arch Pathol Lab Med. 1995;119(2):167–72.

5. Terasaki F, Yoshinaga K. New Guidelines for Diagnosis of Cardiac Sarcoidosis in Japan. Annals of Nuclear Cardiology. 2017;3(1):42–5.

6. Birnie DH, Sauer WH, Bogun F, Cooper JM, Culver DA, Duvernoy CS, et al. HRS expert consensus statement on the diagnosis and management of arrhythmias associated with cardiac sarcoidosis. Heart Rhythm. 2014;11(7):1305–23.

7. Yoon HY, Kim HM, Kim YJ, Song JW. Prevalence and incidence of sarcoidosis in Korea: a nationwide population-based study. Respir Res. 2018;19(1):158.

8. Park JE, Kim YS, Kang MJ, Kim CJ, Han CH, Lee SM, et al. Prevalence, incidence, and mortality of sarcoidosis in Korea, 2003-2015: A nationwide population-based study. Respir Med. 2018;144s:S28–s34.

9. Wu CH, Chung PI, Wu CY, Chen YT, Chiu YW, Chang YT, et al. Comorbid autoimmune diseases in patients with sarcoidosis: A nationwide case-control study in Taiwan. J Dermatol. 2017;44(4):423–30.

10. Pietinalho A, Hiraga Y, Hosoda Y, Löfroos AB, Yamaguchi M, Selroos O. The frequency of sarcoidosis in Finland and Hokkaido, Japan. A comparative epidemiological study. Sarcoidosis. 1995;12(1):61–7.

11. Arkema EV, Grunewald J, Kullberg S, Eklund A, Askling J. Sarcoidosis incidence and prevalence: a nationwide register-based assessment in Sweden. Eur Respir J. 2016;48(6):1690–9.

12. Fidler LM, Balter M, Fisher JH, To T, Stanbrook MB, Gershon A. Epidemiology and health outcomes of sarcoidosis in a universal healthcare population: a cohort study. Eur Respir J. 2019;54(4).

13. Kandolin R, Lehtonen J, Airaksinen J, Vihinen T, Miettinen H, Ylitalo K, et al. Cardiac sarcoidosis: epidemiology, characteristics, and outcome over 25 years in a nationwide study. Circulation. 2015;131(7):624–32.

14. Chapelon-Abric C, de Zuttere D, Duhaut P, Veyssier P, Wechsler B, Huong DLT, et al. Cardiac sarcoidosis: a retrospective study of 41 cases. Medicine (Baltimore). 2004;83(6):315–34.

15. Hulten E, Aslam S, Osborne M, Abbasi S, Bittencourt MS, Blankstein R. Cardiac sarcoidosis-state of the art review. Cardiovasc Diagn Ther. 2016;6(1):50–63.

16. Ipek E, Demirelli S, Ermis E, Inci S. Sarcoidosis and the heart: A review of the literature. Intractable Rare Dis Res. 2015;4(4):170–80.

17. Chiba T, Nakano M, Hasebe Y, Kimura Y, Fukasawa K, Miki K, et al. Prognosis and risk stratification in cardiac sarcoidosis patients with preserved left ventricular ejection fraction. J Cardiol. 2020;75(1):34–41.

18. Nabeta T, Kitai T, Naruse Y, Taniguchi T, Yoshioka K, Tanaka H, et al. Risk stratification of patients with cardiac sarcoidosis: the ILLUMINATE-CS registry. Eur Heart J. 2022;43(36):3450–9.

19. Roberts WC, McAllister HA, Jr., Ferrans VJ. Sarcoidosis of the heart. A clinicopathologic study of 35 necropsy patients (group 1) and review of 78 previously described necropsy patients (group 11). Am J Med. 1977;63(1):86–108.

20. Doughan AR, Williams BR. Cardiac sarcoidosis. Heart. 2006;92(2):282–8.

21. Akashi H, Kato TS, Takayama H, Naka Y, Farr M, Mancini D, et al. Outcome of patients with cardiac sarcoidosis undergoing cardiac transplantation--single-center retrospective analysis. J Cardiol. 2012;60(5):407–10.

22. HCUP Nationwide Readmissions Database (NRD). Healthcare Cost and Utilization Project (HCUP). 2010-2019. Agency for Healthcare Research and Quality, Rockville, MD. Available from: www.hcup-us.ahrq.gov/nrdoverview.jsp.

23. Deyo RA, Cherkin DC, Ciol MA. Adapting a clinical comorbidity index for use with ICD-9-CM administrative databases. J Clin Epidemiol. 1992;45(6):613–9.

24. Patel NJ, Deshmukh A, Pant S, Singh V, Patel N, Arora S, et al. Contemporary trends of hospitalization for atrial fibrillation in the United States, 2000 through 2010: implications for healthcare planning. Circulation. 2014;129(23):2371–9.

25. Rosenthal DG, Fang CD, Groh CA, Nah G, Vittinghoff E, Dewland TA, et al. Heart Failure, Atrioventricular Block, and Ventricular Tachycardia in Sarcoidosis. J Am Heart Assoc. 2021;10(5):e017692.

26. Birnie DH, Kandolin R, Nery PB, Kupari M. Cardiac manifestations of sarcoidosis: diagnosis and management. Eur Heart J. 2017;38(35):2663–70.

27. Murtagh G, Laffin LJ, Beshai JF, Maffessanti F, Bonham CA, Patel AV, et al. Prognosis of Myocardial Damage in Sarcoidosis Patients With Preserved Left Ventricular Ejection Fraction: Risk Stratification Using Cardiovascular Magnetic Resonance. Circ Cardiovasc Imaging. 2016;9(1):e003738.

28. Ziaeian B, Fonarow GC. Epidemiology and aetiology of heart failure. Nat Rev Cardiol. 2016;13(6):368–78.

29. Zhou Y, Lower EE, Li HP, Costea A, Attari M, Baughman RP. Cardiac Sarcoidosis: The Impact of Age and Implanted Devices on Survival. Chest. 2017;151(1):139–48.

30. Kandolin R, Lehtonen J, Graner M, Schildt J, Salmenkivi K, Kivistö SM, et al. Diagnosing isolated cardiac sarcoidosis. J Intern Med. 2011;270(5):461–8.

31. Behnam SM, Behnam SE, Koo JY. TNF-alpha inhibitors and congestive heart failure. Skinmed. 2005;4(6):363–8.

32. Desai R, Kakumani K, Fong HK, Shah B, Zahid D, Zalavadia D, et al. The burden of cardiac arrhythmias in sarcoidosis: a population-based inpatient analysis. Ann Transl Med. 2018;6(17):330.

33. Yazaki Y, Isobe M, Hiroe M, Morimoto S, Hiramitsu S, Nakano T, et al. Prognostic determinants of long-term survival in Japanese patients with cardiac sarcoidosis treated with prednisone. Am J Cardiol. 2001;88(9):1006–10.

34. Kato Y, Morimoto S, Uemura A, Hiramitsu S, Ito T, Hishida H. Efficacy of corticosteroids in sarcoidosis presenting with atrioventricular block. Sarcoidosis Vasc Diffuse Lung Dis. 2003;20(2):133–7.

35. Yodogawa K, Seino Y, Ohara T, Takayama H, Katoh T, Mizuno K. Effect of corticosteroid therapy on ventricular arrhythmias in patients with cardiac sarcoidosis. Ann Noninvasive Electrocardiol. 2011;16(2):140–7.

36. Patel N, Kalra R, Doshi R, Arora H, Bajaj NS, Arora G, et al. Hospitalization Rates, Prevalence of Cardiovascular Manifestations, and Outcomes Associated With Sarcoidosis in the United States. J Am Heart Assoc. 2018;7(2).

37. Nationwide Readmissions Database (NRD), Healthcare cost and utilization project (HCUP), Agency for Healthcare Research and Quality, 2010 – 2019 Available from: https://hcup-us.ahrq.gov/db/nation/nrd/LimitationsonUsingtheNRD.pdf.

38. Murtagh G, Laffin LJ, Patel KV, Patel AV, Bonham CA, Yu Z, et al. Improved detection of myocardial damage in sarcoidosis using longitudinal strain in patients with preserved left ventricular ejection fraction. Echocardiography. 2016;33(9):1344–52.

39. Yasutake H, Seino Y, Kashiwagi M, Honma H, Matsuzaki T, Takano T. Detection of cardiac sarcoidosis using cardiac markers and myocardial integrated backscatter. Int J Cardiol. 2005;102(2):259–68.

